# Tremor asymmetry and the development of bilateral phase-specific deep brain stimulation for postural tremor

**DOI:** 10.1101/2025.01.29.25321310

**Authors:** Shenghong He, Alceste Deli, Timothy O West, Fernando R Plazas, Alek Pogosyan, Christoph Wiest, Laura Wehmeyer, Fahd Baig, Francesca Morgante, Pablo Andrade, Michael G Hart, James J. FitzGerald, Veerle Visser-Vandewalle, Erlick A Pereira, Alexander L Green, Huiling Tan, Hayriye Cagnan

## Abstract

**Background:** Tremor phase-locked deep brain stimulation (DBS) has been shown to modulate symptom severity in patients with postural tremor, including essential (ET) and dystonic tremor (DT). This, provides a potential alternative therapy that targets the underlying pathological oscillations with less energy delivered to the brain than existing systems. Previous phase-locked DBS studies focused on evaluating the effect of unilateral stimulation on the tremor-dominant hand. Considering that postural tremor usually presents bilaterally, it remains unknown how tremor asymmetry interacts with stimulation in the context of bilateral phase-locked DBS, which was investigated in the current study.

**Methods:** Archival limb acceleration from nine ET patients with postural tremor was analysed to explore potential asymmetries in tremor amplitude, frequency, and instability across the two upper limbs, and the relationship between these asymmetries and continuous high-frequency DBS (cDBS). Then, as a proof of concept, bilateral phase-locked DBS was tested in one ET and one DT patient with postural tremor and chronically implanted DBS devices.

**Results:** Our results confirmed that postural tremor is asymmetric; larger tremor power being associated with smaller amplitude and frequency stability in one hand compared to the other. These asymmetries were significantly reduced during cDBS. The effects of cDBS on aforementioned characteristics were greater on the tremor with larger amplitude. Accordingly, the effects of bilateral phasic DBS were also asymmetric.

**Conclusions:** This study creates a better understanding of tremor asymmetry and its relationship with DBS and provides important insights for the development of patient specific approaches for tremor.

## Introduction

Tremor is one of the most common symptoms in movement disorders.^1^ Deep brain stimulation (DBS) targeting the ventral intermediate nucleus (VIM) of the thalamus is a standard treatment for medication refractory tremors.^2^ However, continuous high-frequency DBS (cDBS) may disrupt not only pathological neural signals driving patients’ symptoms, but also physiological neural activities giving rise to stimulation-induced side effects such as impairments in speech, balance, and gait, particularly when the patients adapt to the stimulation over time and thus higher stimulation intensities are required.^2,3^ Previous studies demonstrated that significant tremor relief can be achieved in selected patients during unilateral DBS, phase-locked to tremor from the contralateral hand.^4,5^ Considering that most tremor patients have bilateral tremor,^6^ it remains unknown how postural tremor asymmetry interacts with bilateral phase-locked DBS. In this study, we compared multiple tremor characteristics across both hands in no DBS and cDBS conditions in nine patients with essential tremor (ET). In addition, we piloted bilateral, tremor phase-locked DBS in two tremor patients (1 ET and 1 DT). Our results suggest that postural tremor as well as the effects of both cDBS and tremor phase-locked DBS are asymmetric, indicating that bilateral phase-locked DBS, targeting distinct phases optimized to each side’s tremor would provide superior therapeutic effects.

## Methods

Eleven patients (aged 70 ± 5.77 years, four females) with postural tremor participated in this study (P1-P6 were published previously,^7^ P10 was diagnosed with dystonic tremor (DT) while all other participants were diagnosed with ET). All participants underwent bilateral implantations of DBS electrodes targeting the VIM thalamus and/or posterior subthalamic area (PSA), as shown in Supplementary Figure 1. The study was approved by the local ethics committees and all patients provided their informed written consent according to the Declaration of Helsinki. Recordings from nine patients were conducted 3 to 5 days after the electrode implantation, when the DBS leads were temporarily externalized, and a configurable neurostimulator was used to deliver cDBS. For cases with implanted directional leads, segmented contacts were used in the ring mode. The stimulation contact was selected based on: 1) imaging data and 2) a contact searching procedure to select the final stimulation contact for each hemisphere. To this end, we delivered cDBS initially at 0.5 mA, then progressively increased the amplitude in 0.5 mA increments, until clinical benefit was observed without side-effects such as paraesthesia, or until 3.5 mA was reached as the maximum amplitude. Tremor phase-locked DBS experiments with the remaining two patients were conducted after internalization of the leads and implantable pulse generators (IPGs). Nexus-D4 (Medtronic, MN, USA), an investigational device, was used to deliver tremor-phase-locked bilateral stimulation. We tracked instantaneous tremor phase from both left and right hands using previously described methods.^4,5^ When a desired tremor phase was detected from one of the hands, a transistor-transistor logic (TTL) pulse was sent to Nexus-D4 resulting in a brief burst of stimulation (about 35 ms)^4^ to be delivered to the contralateral VIM (time delay between each TTL and stimulation onset was 109.05 ± 2.42 ms, which was mainly induced by the wireless communication between Nexus-D4 and IPG), at their existing clinical settings (e.g., frequency, amplitude, contact, etc.). Subsequently, a brief burst of stimulation was delivered to the ipsilateral hemisphere with a fixed interval of 71.48 ± 3.18 ms. For instance, if tracking the left hand, Nexus-D4 was used to trigger stimulation to first the right VIM followed by a burst of stimulation to the left VIM. Subsequently, we used tremor phase derived from the other hand to control stimulation delivered to both hemispheres. This allowed us to investigate the effect of tracking tremor phase from one hand to control stimulation timing bilaterally and the effect of tracking tremor phase from both hands through post-hoc analysis. Clinical and stimulation details of all patients are summarised in Supplementary Table I.

Participants were asked to maintain a tremor-provoking posture such as raising both arms to shoulder level with flexed elbows and the fingers of both hands pointing to the centre, while, limb acceleration was acquired using triaxial accelerometers taped to the back of both hands, and simultaneously recorded using a TMSi Porti amplifier (TMS International) at a sampling rate of 2048 □ Hz (P1-P9) or using a CED Power1401-3A data acquisition interface (Cambridge Electronic Design, Cambridge, UK) at a sampling rate of 10 KHz (P10-P11). To investigate tremor asymmetry, we (1) segmented limb acceleration measurements into 2-s non-overlapping trials, (2) quantified tremor characteristics including frequency, power, and cycle-by-cycle instability defined by the standard deviations of amplitude and frequency across tremor cycles for each 2-s trial, and (3) compared them between hands in both no DBS and cDBS conditions, as shown in Fig. 1A. To investigate the effect of tremor phase-locked stimulation, for each phasic stimulation trial (here one trial represents tracking a specific tremor phase from one of the limbs for 7 s), we quantified the change in tremor severity on one hand or both hands by comparing the tremor amplitude between the last 1-second of phasic stimulation and the last 1-second before stimulation onset at that specific phase (Fig. 2A), similar to the method used in previous studies.^4,5^

**Figure 1.**
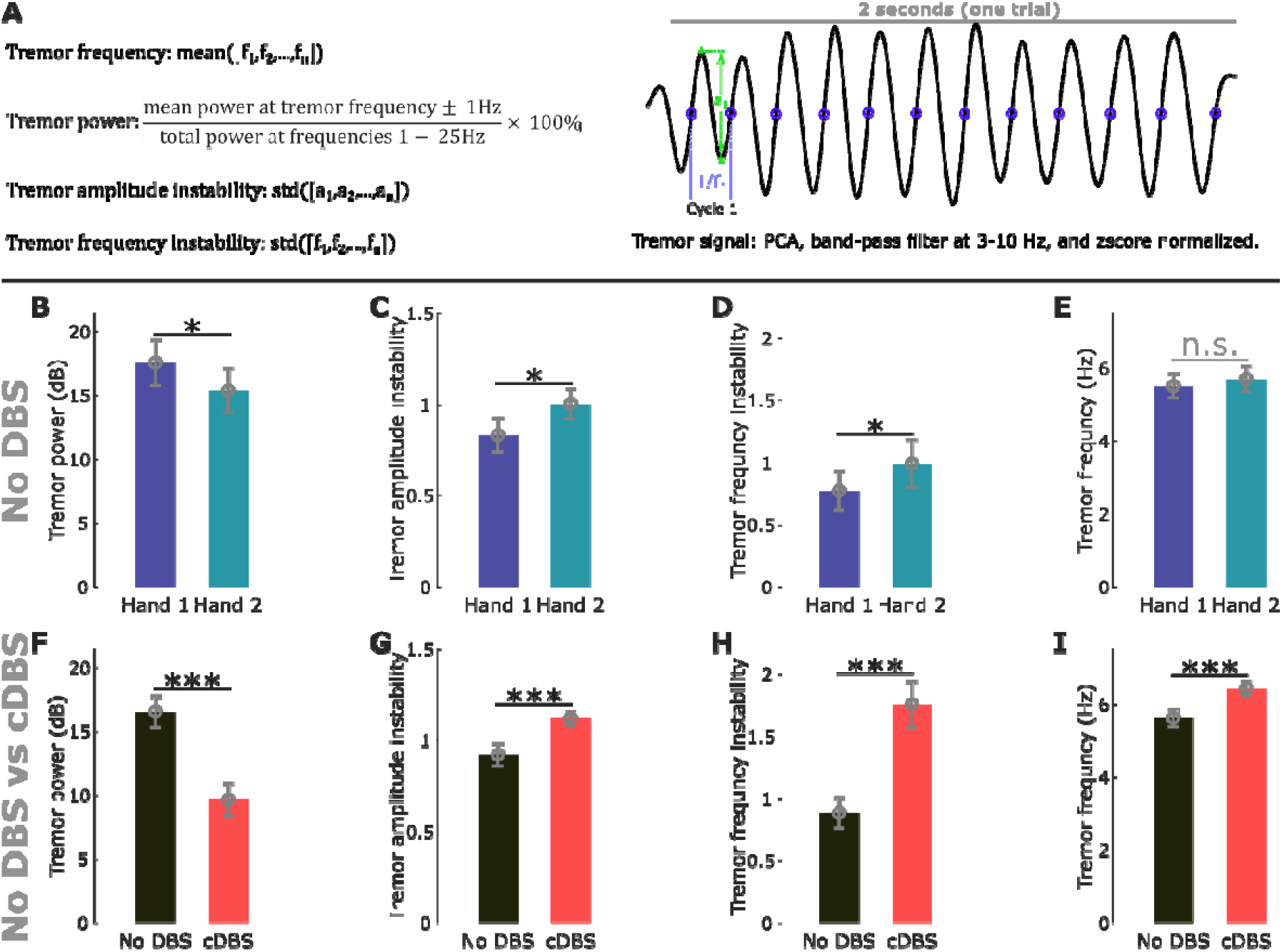
Postural tremor was asymmetric in terms of power and instability. These asymmetric measurements were modulated differently by cDBS on different hands. **(A)** A demonstration of the quantifications of tremor frequency, power, amplitude instability, and frequency instability. **(B)-(E)** Comparisons of tremor power (B), amplitude instability (C), frequency instability (D), and frequency (E) between tremor pre-dominant (Hand 1) and non-dominant (Hand 2) hands measured during no DBS condition. **(F)-(I)** Comparisons of tremor power (F), amplitude instability (G), frequency instability (H), and frequency (I) between no DBS and cDBS conditions.

**Figure 2.**
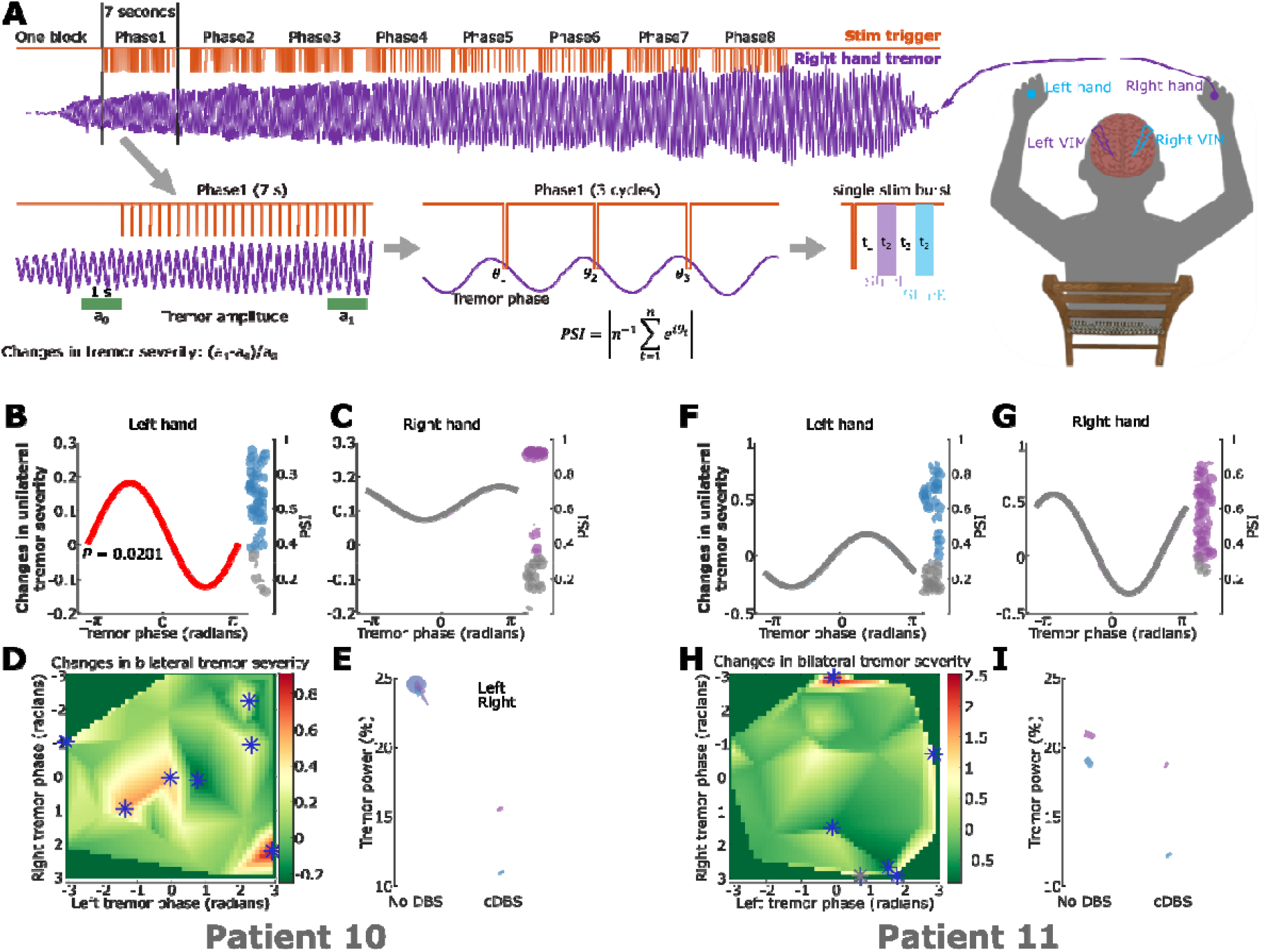
Protocol of tremor phase locked DBS and its effect on tremor severity. **(A) A demonstration of right-hand tremor phase-locked bilateral stimulation.** Tremor signals are recorded using a triaxial accelerometer. The dominant tremor axis is determined and real-time filtered at the patient specific tremor frequency band, as shown in purple. Stimulation is delivered in blocks, with each block containing eight trials. In each trial, stimulation is triggered at one of the eight predefined tremor phases for 7 seconds, as shown in orange. Within each trial, one trigger is sent out at a specific tremor phase. Following each trigger, two bursts of stimulation with a fixed interval of 71.48 ± 3.18 ms (t_3_) were delivered to the contralateral followed by the ipsilateral VIM relative to the hand used for tremor phase tracking. A fixed time interval of 109.05 ± 2.42 ms (t_1_) was induced by the Nexus-D4 between each trigger and the first burst of stimulation. The duration of each burst of stimulation was about 35 ms (t_2_). Stimulation effect is evaluated by comparing the tremor amplitude (on one or both hands) in the last second within each trial relative to one second immediately before the start of each trial, i.e., changes in tremor severity. The accuracy of phase-locking is evaluated by quantifying the phase synchrony index (PSI) across all stimulation triggers within each trial. **(B)-(C)** Effects of bilateral tremor phase-locked stimulation on one hand. Each bar indicates the effect of a specific tremor phase bin, with the curve representing the fitted sine curve considering all bins. Each dot on the right side indicates accuracy of the corresponding phase-locked stimulation trial, quantified using PSI. Red sine curve indicates significance against the surrogate distribution after correcting for multiple comparisons. Grey dots for PSI indicate non-significant phase-locking trials against surrogate distributions, which have been excluded from the analysis. **(D)** Effects of bilateral tremor phase-locked stimulation on both hands. Bilateral phasic stimulation effect on tremor severity measured from both hands. Blue stars indicate marginally significant phasic effects against surrogate distribution, which did not survive multiple comparison corrections though. **(E)** Effects of cDBS on tremor severity for left (blue) and right (purple) hands. **(F)-(I)** The same as (B)-(E) but for patient 11.

Statistical analyses were conducted using custom-written scripts in MATLAB R2021-b (The MathWorks Inc, Nantucket, Massachusetts). Tremor characteristics were quantified on individual trial basis (including frequency, power, amplitude instability, and frequency instability), and a generalized linear mixed effect modelling (GLME) was used to investigate the difference between hands, stimulation conditions, as well as the interaction between the two.^8^ Multiple comparisons applied to these measurements were corrected using false discovery rate (FDR) approach (Benjamini and Hochberg, 1995; Benjamini and Yekutieli, 2001).^9,10^ The estimated value with standard error of the coefficient (*k* ± SE), pre-corrected P-values as well as their significances after FDR correction were reported. To test the effects of bilateral phasic stimulation, we tested significance of the change in tremor severity for each phase bin against a surrogate distribution (D_1_) with 1,000,000 points representing the natural tremor variability without DBS. Specifically, we randomly selected 50,000 7-s segments of tremor during no DBS and quantified the natural changes in tremor severity to generate a surrogate distribution D_0_. For each phase bin, we randomly selected N values from D_0_ and quantified their median value (N corresponding to the number of stimulation trials delivered at this phase bin). This was repeated 1,000,000 times leading to the surrogate distribution D_1_.^4,5^ To investigate the overall tremor modulation and different modulation patterns across hands, we fitted a sine curve to the changes in tremor severity over all tested tremor phase bins for each hand. The amplitude of the fitted sine curve was tested against a surrogate distribution (D_2_) with 1,000,000 points, in which each point indicated the amplitude of the fitted sine curve derived from the surrogate distribution D_1_ for the same hand.^11^ To control for the accuracy of phase-locked stimulation, we quantified the phase synchrony index (PSI) across tremor phases, at which stimulation was delivered in each trial, and tested this against a surrogate distribution with 50,000 points, with each point indicating the PSI quantified after shuffling the tremor phase relative to the stimulation timing.

### Data availability

The data and codes will be shared upon request from the corresponding author.

## Results

### 1. Postural tremor was asymmetric across hands

In the absence of stimulation, on average, the tremor-dominant hand (“Hand 1”) defined as the hand with larger tremor power (Fig. 1B, *k* = -2.4217 ± 1.0275, *P* = 0.0185), had significantly less tremor instability in terms of amplitude (Fig. 1C, *k* = 0.1492 ± 0.0684, *P* = 0.0293) and frequency (Fig. 1D, *k* = 0.2362 ± 0.1067, *P* = 0.0270) when compared with the non-tremor-dominant hand (“Hand 2”), although the peak tremor frequencies were similar between the hands (Fig. 1E). These differences were no longer significant during cDBS (Supplementary Table II), i.e., cDBS reduced tremor asymmetries. Overall, apart from reducing tremor power (Fig. 1F, *k* = -7.0614 ± 1.2131, *P =* 6.379 × 10^-9^), cDBS significantly increased tremor instability (amplitude: Fig. 1G, *k* = 0.2129 ± 0.0634, *P =* 0.0008; frequency: Fig. 1H, *k* = 0.8997 ± 0.1529, *P* = 4.3655 × 10^-9^) and peak tremor frequency (Fig. 1I, *k* = 0.8026 ± 1.1518, *P =* 1.3082 × 10^-7^). An interaction analysis (Supplementary Table II) revealed that the tremor asymmetries, i.e., the differences in the tremor characteristics between Hand 1 and Hand 2, were significantly reduced by cDBS (except for frequency instability), and the effects of cDBS on these tremor characteristics were significantly stronger on Hand 1 compared to Hand 2 (except for frequency instability).

### 2. Phasic DBS has different effects on different hands

For both patients and hemispheres, while considering the changes in tremor severity on one hand only, when bilateral phasic stimulation was delivered to the contralateral VIM first, there was no distinct phase bin that showed significant modulation compared with the natural tremor variability (see the statistical analysis part in Methods for more details). However, the overall stimulation effect was significant in P10, left hand (Fig. 2B, *P* = 0.0201 against surrogate distribution) when using a fitted sinusoid to test significance and using FDA for multiple comparisons correction. This was reflected during cDBS as well: the most prominent effect was seen in P10 left hand (Fig. 2E and I). Tremor modulation profiles during phasic DBS (Fig. 2B and C, F and G), and the modulation of other tremor characteristics with cDBS (Supplementary Fig. 2) were different across hands, analogous to the results highlighted in Part 1. While considering the changes in tremor severity for both left and right hands (by taking the average change in tremor severity) for a given tremor phase combination derived from instantaneous tremor phases from both hands, both patients showed multiple phase combinations that provided marginally significant effects on bilateral tremor severity (Fig. 2D and H, blue stars, *P* < 0.05 against surrogate distribution, although none of them survived multiple comparison corrections). These results suggested that phase-locked DBS delivered to the two hemispheres targeting the optimal tremor phases should be determined for left- and right-hand jointly.

## Discussion

To limit the emergence of stimulation induced side-effects, different closed-loop DBS protocols have been proposed, including switching on DBS only when tremor or a tremor provoking movement is detected^7,12,13^ or delivering stimulation time-locked to a specific tremor rhythm^4,5^. The first strategy is based on the intermittent nature of postural tremor in ET and DT, while the latter approach aims to specifically disrupt the underlying neural activity related to tremor. In a recent study involving a clinical survey of 487 individuals diagnosed with ET, Whaley et al. reported that about half (52%) of the cohort reported bilateral tremor onset, and about 90% of the individuals eventually presented bilateral tremor.^6^ A separate study found that while bilateral VIM DBS provided greater overall tremor reduction across both sides compared to unilateral DBS, unilateral stimulation was just as effective in alleviating tremors in the contralateral hand.^14^ Here, we showed that postural tremor is asymmetric in terms of cycle-by-cycle tremor frequency, power, and instability (amplitude and frequency) in the absence of DBS as well as when bilateral cDBS was on. Effects of phase-locked DBS on different hands were also asymmetric. Results from a separate study also showed that the efferent thalamic to tremor connectivity was lateralized in ET, with a stronger connectivity from the contralateral VIM, which was associated with bigger and more stable tremor as well as larger cDBS effects. In addition, more unstable tremor was associated with stronger cross-hemisphere coupling between left and right thalami.^15^ These results together suggest that tremor in different hands might be associated with different but interacting oscillatory sources. Bilateral phase-locked DBS independently targeting the optimal suppressive phases for left- and right-hand tremor might work better in disrupting the relevant oscillatory sources, and thus be more effective at suppressing pathological tremors, whist simultaneously reducing the total electrical energy delivered.

## Data Availability

The data and codes will be shared upon request from the corresponding author.

## Acknowledgement

This work was supported by the Medical Research Council (MR/R020418/1, MR/X023141/1, and MC_UU_00003/2) and the Guarantors of Brain. We thank the participating patients for making this study possible and thank Medtronic for providing the investigational device Nexus-D4 used in this study.

## References

1. Louis ED, Ferreira JJ. How common is the most common adult movement disorder? Update on the worldwide prevalence of essential tremor. Movement Disorders. 2010 Apr 15;25(5):534–41.

2. Baizabal-Carvallo JF, Kagnoff MN, Jimenez-Shahed J, Fekete R, Jankovic J. The safety and efficacy of thalamic deep brain stimulation in essential tremor: 10 years and beyond. Journal of Neurology, Neurosurgery & Psychiatry. 2014 May 1;85(5):567–72.

3. Zhang K, Bhatia S, Oh MY, Cohen D, Angle C, Whiting D. Long-term results of thalamic deep brain stimulation for essential tremor. Journal of neurosurgery. 2010 Jun 1;112(6):1271–6.

4. Cagnan H, Pedrosa D, Little S, Pogosyan A, Cheeran B, Aziz T, Green A, Fitzgerald J, Foltynie T, Limousin P, Zrinzo L. Stimulating at the right time: phase-specific deep brain stimulation. Brain. 2017 Jan 1;140(1):132–45.

5. Reis C, He S, Pogosyan A, Haliasos N, Low HL, Misbahuddin A, Aziz T, Fitzgerald J, Green AL, Denison T, Cagnan H. Phase-specific Deep Brain Stimulation revisited: effects of stimulation on postural and kinetic tremor. medRxiv. 2022 Jun 21:2022–06.

6. Whaley NR, Putzke JD, Baba Y, Wszolek ZK, Uitti RJ. Essential tremor: phenotypic expression in a clinical cohort. Parkinsonism & related disorders. 2007 Aug 1;13(6):333–9.

7. He S, Baig F, Mostofi A, Pogosyan A, Debarros J, Green AL, Aziz TZ, Pereira E, Brown P, Tan H. Closed-Loop Deep Brain Stimulation for Essential Tremor Based on Thalamic Local Field Potentials. Movement Disorders. 2021 Apr;36(4):863–73.

8. Lo S, Andrews S. To transform or not to transform: Using generalized linear mixed models to analyse reaction time data. Frontiers in psychology. 2015 Aug 7;6:1171.

9. Benjamini Y, Hochberg Y. Controlling the false discovery rate: a practical and powerful approach to multiple testing. Journal of the Royal statistical society: series B (Methodological). 1995 Jan;57(1):289–300.

10. Benjamini Y, Yekutieli D. The control of the false discovery rate in multiple testing under dependency. Annals of statistics. 2001 Aug 1:1165–88.

11. van Es MW, Marshall TR, Spaak E, Jensen O, Schoffelen JM. Phasic modulation of visual representations during sustained attention. European Journal of Neuroscience. 2022 Jun; 55(11-12):3191–208.

12. Herron JA, Thompson MC, Brown T, Chizeck HJ, Ojemann JG, Ko AL. Chronic electrocorticography for sensing movement intention and closed-loop deep brain stimulation with wearable sensors in an essential tremor patient. Journal of neurosurgery. 2016 Nov 18;127(3):580–7.

13. Opri E, Cernera S, Molina R, Eisinger RS, Cagle JN, Almeida L, Denison T, Okun MS, Foote KD, Gunduz A. Chronic embedded cortico-thalamic closed-loop deep brain stimulation for the treatment of essential tremor. Science translational medicine. 2020 Dec 2;12(572):eaay7680.

14. Huss DS, Dallapiazza RF, Shah BB, Harrison MB, Diamond J, Elias WJ. Functional assessment and quality of life in essential tremor with bilateral or unilateral DBS and focused ultrasound thalamotomy. Movement Disorders. 2015 Dec;30(14):1937–43.

15. He S, West TO, Plazas FR, Wehmeyer L, Pogosyan A, Deli A, Wiest C, Herz DM, Simpson T, Andrade P, Baig F. Cortico-thalamic tremor circuits and their associations with deep brain stimulation effects in essential tremor. Brain. 2024 Nov 27:awae387.

